# Identifying Facilitators of and Barriers to Digital Health Literacy in Pediatric Rheumatic Diseases: A Scoping Review Protocol

**DOI:** 10.1101/2025.09.16.25335907

**Authors:** Craig Eling, Alan M. Rosenberg, Jennifer N. Stinson, Maryam Mehtar, Jasmin Bhawra, Mary Chipanshi, Roona Sinha, Donna Goodridge

## Abstract

**Background:** Digital health literacy (DHL) is a set of skills needed to positively integrate health information acquired from digital sources into health behaviors and lifestyles. Digital health is defined as the use of technology to deliver, manage, discuss, and improve health. Higher levels of DHL are associated with better health-related outcomes and disease management, while lower levels of DHL serve as a barrier to improving health. In the context of childhood rheumatic diseases, fostering robust DHL can empower children, adolescents, and their primary caregivers to navigate the complexities of managing and understanding health information in this digital era. The objective of this report is to develop a protocol that can be used to identify existing assessments of DHL and knowledge about factors that promote or impede DHL in children and adolescents with chronic rheumatic diseases and their primary caregivers.

**Methods:** The scoping review protocol applies the JBI scoping review framework and the Preferred Reporting Items for Systematic Review and Meta-Analysis extension for scoping reviews. The protocol will identify full-text English language studies and dissertations published after 1974 that identify facilitators of and barriers to DHL in patients younger than age 19-years diagnosed with a chronic rheumatic disease and/or their primary caregivers. In this protocol qualitative, quantitative, and mixed methods studies are eligible for inclusion with no limitation on the geographical origins of the publications; grey literature will be excluded. The protocol requires two researchers to independently screen titles and abstracts for inclusion, followed by a full text review for data extraction. Descriptive statistics around identified facilitators of and barriers to DHL, populations studied, and other pertinent data will be provided. Qualitative data will be open coded and categorized around the research questions. A narrative summary will accompany the results.

**Discussion:** The novel and comprehensive search strategy outlined by this protocol will enable ongoing research related to the interaction of technology, the healthcare system, and individuals affected by having a pediatric rheumatic disease.

**Scoping review registration:** The protocol has been preregistered on the Open Science Framework (https://osf.io/g4jbh/).

## Background

Digital health refers to the use of digital technology to manage and improve health (1,2). Digital health includes telehealth, telemedicine, virtual medicine, mobile health (mHealth), information technology associated with healthcare, wearable devices, and emerging technologies. Digital health is delivered through many platforms, applications, websites, and sensors, and may utilize artificial intelligence and virtual or augmented reality (1).

Pediatric rheumatic diseases (PRD) are a class of chronic systemic inflammatory disorders that include an array of musculoskeletal and connective tissue disorders, with juvenile idiopathic arthritis (JIA) being the most common (3). PRD can have adverse effects on growth and development, quality of life, academic performance, mental health, and transitioning to adult health care (4).

Digital health holds particular promise for improving both health outcomes and quality of life for young people and families affected by PRD through the use of technologies that provide reassurance and enhanced ability to self-manage diseases (5). Access to digital health resources can contribute to enhancing disease education and management, connections with patient and parent support groups, fostering alliances with advocacy and educational associations, and use of digital communication methods with health care providers (6,7). Promoting and facilitating digital health literacy among children and families affected by having a PRD can help navigate and manage the complexities of their conditions.

To utilize and benefit from digital health, users must have a prerequisite level of digital health literacy (DHL) (8–10). DHL is a set of skills that allows digital health users to seek, access, appraise, utilize, and integrate health information beneficially into their daily routines, communicate about health, and gain benefit from apps and digital services (8–10). Higher DHL aids in accessing, finding, and understanding health information, navigating the complexities of the healthcare system, interpreting disease status, monitoring treatments, and having electronic communication with health care providers (4,11,12). This results in better disease management in children and adolescents with chronic childhood diseases, including PRDs, and their primary caregivers (4,10–12). Conversely, low DHL is associated with poor health outcomes and the amplification of pre-existing health and social disparities (7,9,13). Low DHL can lead to barriers in access to care, less benefit from treatments, poorer outcomes, fear of disruption in care, increased isolation, inability to find reliable health information, and confusion about the integrity of online health information (7,9,13).

Digital health can provide more efficient, timely, cost-effective, and convenient health care services, allowing people to have greater access to health-related services and personal health data that has previously been unaffordable and unavailable (1,9,14). Digital health has the potential to address and correct healthcare inequalities based on gender, disabilities, living in rural and remote settings, and mental health issues (1,14,15). Specific to PRD, digital health can provide a solution to the inadequate access to pediatric rheumatologists and avoid travel to and from pediatric rheumatic services that are increasingly being centralized at children’s hospitals (4). Furthermore, the increased uptake and migration to digital health was greatly accentuated by the COVID-19 pandemic (6,7,13). In fact, the COVID-19 pandemic caused an “infodemic” of health information that varied in its validity and spread “faster than the coronavirus itself” (7). Thus, DHL has been described as the “social vaccine” for the infodemic (9).

### Rationale and Objectives

The rationale for this ScR protocol stems from the recognition of the limited awareness and information about DHL in the PRD population. A broad search of the literature focusing on children/adolescents with chronic disease, parents of children/adolescents with chronic disease, parents of healthy children/adolescents, and adults with rheumatic disease did not reveal any results specific to PRD. The non-PRD results of this search are presented in Appendix 1 and were used to generate the objectives and research questions for this ScR protocol. This ScR protocol intends to provide guidance for undertaking a ScR to answer two questions: “what are the facilitators of and barriers to DHL identified in the literature for children and adolescents with PRD and/or primary caregivers of children and adolescents with PRD?” This ScR will help to collate and appraise current knowledge regarding DHL in the PRD population, describe the nature and methodologies of extant research, and identify any gaps in the research (29).

### Methodology

This protocol was written as per the JBI ScR framework (30) and in accordance with PRISMA-ScR guidelines (31). This protocol was registered with the Open Science Framework (https://osf.io/g4jbh/). This scoping review (ScR) protocol incorporates the application Covidence (32), a browser-based tool that streamlines the review process, allowing researchers conducting systematic reviews to effectively collaborate and efficiently organize, screen, and extract data from studies. Covidence complies with Preferred Reporting Items for Systematic Review and Meta-Analysis extension for scoping reviews (PRISMA-ScR), can assess risk of bias in individual studies, and can present real-time statistics comparing approval and denial rates among reviewers. NVivo qualitative data software (33) will be used to manage and analyze data. NVivo is a tool that allows researchers to collaborate in the extraction, organization, and coding of both qualitative and quantitative data, including text, video, audio, pictures, graphs, charts, mind maps, and any other non-numerical data formats.

Table 1 shows the population, concept, and context (30) used to develop the research questions used in this proposal. *Facilitators* are defined as any personal, population-level, or systemic factors that develop or promote an individual’s DHL. Conversely, *barriers* are personal, population-level, or systemic factors that delay or weaken an individual’s DHL.

**Table 1.** Population, concept, and context (PCC) framework.

|  |  |
| --- | --- |
| P: Population | Children (aged 0- to 9-years) and adolescents (aged 10- to 19-years) with PRD. Primary caregivers of children or adolescents with PRD. |
| C: Concept | Facilitators to and barriers of DHL. |
| C: Context | Identification of factors serving as facilitators of or barriers to DHL that can affect the delivery of pediatric rheumatic disease digital health-based services. This includes virtual clinic visits, digital communication, educational supports, involvement in online communities, recommendation of specific apps, and other Internet-based resources. |

Primary caregivers are defined as a person or people who chiefly care for the child or adolescent with PRD in their home. This includes helping with daily tasks, providing necessities, and participating day-to-day in actively managing the child’s or adolescent’s PRD. This is traditionally the parental role, although other individuals may fulfill this role, regardless of biological relationship and traditional roles.

#### Identify relevant studies

The electronic databases to be searched include MEDLINE, CINAHL, Embase, Web of Science, and ProQuest Dissertations & Theses Global. The search terms were developed by a librarian (MC) and experts in the subject area (AR, JS, DE, and CE) and piloted through a search of MEDLINE. Prior to finalizing the search terms for this protocol, the draft search terms underwent a review by an independent librarian experienced with systematic reviews, using the Peer Review of Electronic Search Strategies (PRESS) guidelines (34). The feedback from the PRESS review was incorporated into the search strategy, which is presented in Table 2.

**Table 2.** MEDLINE pilot search (Ovid MEDLINE(R) and In-Process, In-Data-Review and Other Non-Indexed Citations and Daily <1974 to January 16, 2024>).

|  |  |  |
| --- | --- | --- |
| # | Query | Results from January 16, 2024 |
| 1 | Literacy/ or Health Literacy/ or Information Literacy/ or Computer Literacy/ or (((("digital health litera*" or "digital health care litera*" or "digital medic* litera*" or "e?health litera*" or "e?healthcare litera*" or "e?care litera*" or "m?health litera*" or "mobil* health litera*" or "m?health?care litera*" or "virtual?health litera*" or "virtual healthcare litera*" or health) adj2 litera*) or litera* or literate or (digital or virtual or e or m)) adj3 litera*).mp. | 1,187,840 |
| 2 | exp Telemedicine/ or exp Telecommunications/ or ("digital health" or "digital health?care" or "digital medicine" or "e?Health" or "e?Health?care" or "e?medicine" or "tele?health" or "tele?health?care" or "tele?medicine" or "m?Health" or "m?Health?care" or "mobile health" or "mobile health?care" or "mobile medicine" or "online health" or "online health?care" or "online medicine" or "mobile app*" or "digital techn*" or "interactive program*" or internet or "web?based" or "communication technolog*" or "digital technolog*" or internet or online or web or mobile or phone or digital or electronic or mobile or app or apps or "information technology" or "Internet technology" or "artificial intelligence" or "wearable technology" or robotic or robot or robotics or "augmented reality" or "virtual reality" or sensors).mp. | 1,494,631 |
| 3 | 1 AND 2 | 133,331 |
| 4 | exp Arthritis/ or ("juvenile idiopathic arth*" or "juvenile chronic arth*" or "juvenile rheumatoid arth*" or "p?ediatric rheum*" or pcJIA or JIA | 322,327 |
|  | or JA or JRA or "childhood arth*" or JCA or "Reiter's Syndrome" or "enthesitis related arth*" or "psoriatic arth*" or "juvenile onset still disease" or "juvenile onset still's disease" or "juvenile oligoarthritis" or "juvenile systemic arth*" or "inflammatory polyarthritis" or "rheumatic arth*").mp. or ((polyarticular or polyarthritis) adj3 juvenile).mp. |  |
| 5 | exp Lupus Erythematosus, Systemic/ or ("systemic lupus erythematosus" or "lupus erythematosus" or "systemic lupus" or SLE or lupus or "childhood systemic lupus" or "p?ediatric systemic lupus" or "childhood SLE" or "p?ediatric SLE" or "p?ediatric?onset systemic lupus" or "childhood?onset systemic lupus" or "childhood?onset SLE" or "p?ediatric?onset SLE" or cSLE or "juvenile?onset systemic lupus" or "juvenile?onset SLE" or neurolupus or "neuro SLE" or "neuropsychiatric systemic lupus" or NPSLE or "CNS lupus" or "central nervous?system lupus" or "central nervous?system SLE").mp. | 106,003 |
| 6 | exp Vasculitis/ or (vasculitis or arteritis or endarteritis or polyarteritis or "Takayasu arteritis" or Kawasaki or "Kawasaki disease" or "IgA vasculitis" or "Schoenlein?henoch purpura" or "Henoch?schoenlein purpura" or "Behcet*" or Granulomatosis or EGPA or GPA or "Churg?Strauss" or "Buerger*" or "Henoch?schoenlein" or "Schoenlein?henoch" or Wegener* or Goodpasture* or "thromboangiitis obliterans" or polyangi* or cryoglobulinemia or Kawasaki* or sarcoidosis or "macrophage activation syndrome" or "anti?glomerular basement membrane" or "anti?GBM" or (ANCA adj2 vasculi*) or (anti?neutrophil adj2 vasculi*) or (immune adj2 vasculi*)).mp. | 188,877 |
| 7 | ("juvenile ankylosing spondylitis" or "enthesitis related arthritis" or sarcoidosis or "macrophage activation syndrome" or "ankylosing spondylarthri*" or "bechterew* disease" or "marie?strumpell disease" or "rheum* spondylitis" or "spondyloarthritis ankylopoietica").mp. | 37,274 |
| 8 | exp Rheumatic Diseases/ or exp Autoimmune Diseases/ or Rheumatic Fever/ or (rheumatism or "rheum* disease*" or "acute rheum* fever*" or "inflammatory rheum*" or "p?ediatric rheum* disease" or "child* rheumatic disease").mp. | 686,341 |
| 9 | 4 OR 5 OR 6 OR 7 OR 8 | 943,662 |
| 10 | exp Child/ or Adolescent/ or exp Infant/ or (p?ediatric* or Infant or child* or adolescent* or newborn or baby or babies or infant or adolescent* or teen* or "school?aged" or preschool* or toddler or kid* or youth* or boy* or girl* or CYP or "Children and young people" or preschool* or "pre?school*" or "play?school" or "school?child*" or school?age* or "pre?teen" or teen* or "young?person*" or "young?people").mp. | 5,651,502 |
| 11 | exp Family/ or exp Family Relations/ or exp Parent Child Relations/ or (parent* or carer* or "care?giver" or carer* or father* or mother* or famil* or "grand?parent*" or "grand?father*" or "grand?mother*?" or guardian* or relativ* or "children?caring" or "families?caring").mp. or (famil* adj2 support).mp. | 3,773,007 |
| 12 | 10 OR 11 | 8,377,659 |
| 13 | 3 AND 9 AND 12 | 770 |
| 14 | exp Animals/ NOT exp Humans/ | 5,185,986 |
| 15 | 13 NOT 14 | 764 |
| 16 | limit 15 to English language | 733 |
| 17 | Limit 16 to yr="1974 -Current" | 733 |

The search utilizes both medical subject headings (MeSH) and keyword searches, combined using the Boolean terms “AND”, “OR”, and “NOT”. The search strategy involves three parts: 1. identifying studies focusing on DHL, using the many synonyms and associated terms; 2. identifying studies focusing on rheumatic conditions, both general and pediatric specific; and 3. identifying studies that focus on children, adolescents, and primary caregivers. These three parts will then be combined for the results that are specific to the facilitators of and barriers to DHL in children or adolescents with PRD and their primary caregivers.

As the search progresses and moves to additional databases, the search strategy will be updated, if necessary, to best address the objective and research questions. Reference lists of the included studies will be hand searched for any applicable articles that were not previously captured.

#### Inclusion Criteria

This ScR protocol will capture published studies meeting the following requirements:

- All qualitative, quantitative, and mixed methods study designs from any discipline of healthcare or clinical setting; systematic reviews that meet the inclusion criteria will also be considered,
- The study’s target population is children (aged 0 to 9 years) and/or adolescents (aged 10 to 19 years) with PRD or primary caregivers of children/adolescents with PRD,
- English language full text, and
- Articles published since January 1, 1974, which is the year that the term health literacy was first used (35).

#### Exclusion Criteria

This ScR protocol will exclude the following:

- Opinion papers, conference proceedings, and editorials,
- Grey literature,
- Studies from complementary and alternative health fields (as examples: chiropractic, naturopathic, and acupuncturist),
- Articles with full text published in a language other than English, and Full-text articles that cannot be accessed either digitally or paper-based.

#### Selection Process

Articles identified by the search strategy defined in this protocol will be imported into Covidence (32) and duplicates will be removed. Two reviewers who will be conducting the article screening will trial the a priori inclusion and exclusion criteria for 20 articles, to finalize these criteria and minimize selection bias (36). Following this, titles and abstracts will be screened for inclusion; a full-text screen will be completed if the article’s eligibility cannot be determined through the title and abstract. Any disagreements in eligibility will be resolved through a third reviewer. This decision process and the results of the search will be documented in a PRISMA-ScR flow diagram (31,37) (Figure 1).

**Fig 1.**
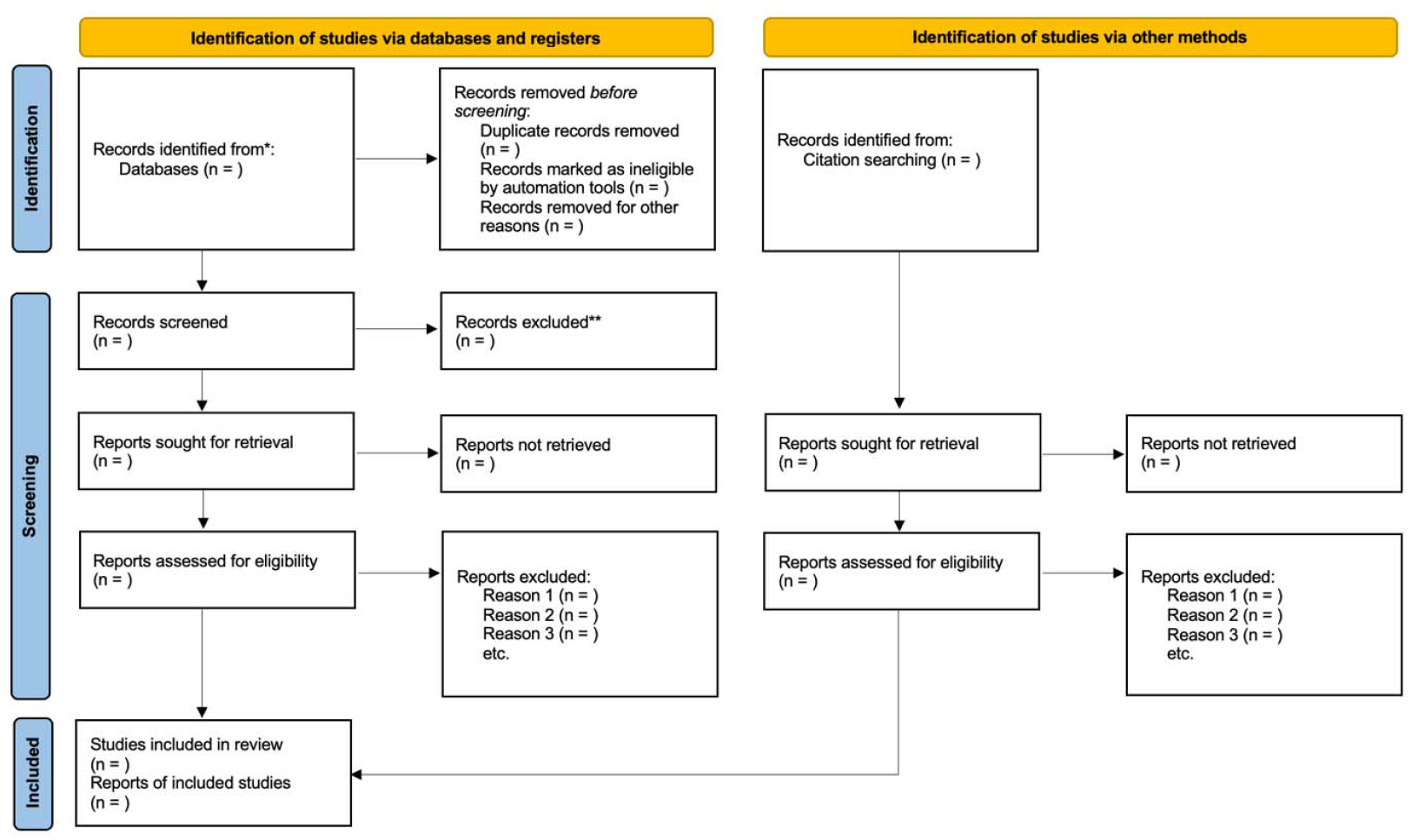
PRISMA flow diagram (37). *Consider, if feasible to do so, reporting the number of records identified from each database or register searched (rather than the total number across all databases/registers). **If automation tools were used, indicate how many records were excluded by a human and how many were excluded by automation tools. From: Page MJ, Moher D, Bossuyt PM, Boutron I, Hoffmann TC, Mulrow CD, Shamseer L, Tetzlaff JM, Akl EA, Brennan SE, Chou R, Glanville J, Grimshaw JM, Hróbjartsson A, Lalu MM, Li T, Loder EW, Mayo-Wilson E, McDonald S, McGuinness LA, Stewart LA, Thomas J, Tricco AC, Welch VA, Whiting P, McKenzie JE. PRISMA 2020 explanation and elaboration: updated guidance and exemplars for reporting systematic reviews. BMJ 2021;372:n160. doi: 10.1136/bmj.n160. For more information, visit: http://www.prisma-statement.org/.

#### Quality Appraisal

All studies included after the full-text review in the ScR will be analyzed with the Mixed Methods Appraisal Tool (MMAT) (38). The MMAT allows researchers to critically appraise studies that are qualitative and randomized controlled trials, nonrandomized, descriptive, and mixed methods studies, but not reviews articles. The tool consists of a checklist where negative responses are used to guide a more thorough review; researchers are discouraged from calculating an overall score for each study. If studies are discovered to be of low-quality, researchers are advised not to exclude them from the results. Rather, researchers are encouraged to contrast the quality of the study with the results, discussing the reasoning and implications of for the low-quality rating (38). Two reviewers will conduct independent analyses of the included studies identified through the application of this protocol using the MMAT.

#### Charting the Data

A data extraction tool based on the JBI extraction instrument template (30) (Table 3) will be developed using an Microsoft Excel spreadsheet (39). The tool will be trialed by the two reviewers The tool may be updated iteratively as the ScR progresses. Data from studies that have passed the screening process will be extracted as per the tool, which will be trialed for five articles by two reviewers prior to acceptance. Once accepted, both reviewers will independently proceed with the data extraction from the included studies.

**Table 3.** Data extraction tool, based on the JBI template (30).

|  |  |
| --- | --- |
| **Identifying Facilitators of and Barriers to Digital Health Literacy in Chronic Childhood Rheumatic Disease** |  |
| ScR Objective | From the literature identify facilitators of and barriers to DHL in young people with chronic rheumatic diseases and their primary caregivers |
| ScR Question | What are the facilitators of and barriers to DHL identified in the literature on children and adolescents with PRD and primary caregivers of children or adolescents with PRD? |
| **Inclusion/Exclusion Criteria** |  |
| Population | Children (aged 0 to 9 years) and adolescents (aged 10 to 19 years) with PRD; primary caregivers of children or adolescents with PRD |
| Concept | Identified facilitators to and barriers of DHL |
| Context | Factors affecting implementation or ongoing use of digital health services in pediatric rheumatology |
| Sources of evidence | Qualitative, quantitative, and mixed methods studies published since 1974 |
| **Evidence source Details and Characteristics** |  |
| Citation details | (author/s, date, title, journal, volume, issue, pages) |
| Country |  |
| Context | (study details pertaining to the objectives and questions, health discipline, health care setting) |
| Participants | (disease/diagnosis, children/youth/primary caregivers) |
| Study type | (qualitative, quantitative, mixed methods, systemic review) |
| **Details extracted from source of evidence** |  |
| Facilitators of DHL | (factors promoting and supporting DHL, including any qualitative themes) |
| Barriers to DHL | (factors delaying or detracting from DHL, including any qualitative themes) |

#### Collating, summarizing, and reporting the results

For the qualitative component of this scoping review protocol, included studies will be analyzed to identify and synthesize key themes, patterns, and insights from the included studies. textual data will be entered into NVivo (33), which will assist in the organization and coding of nodes, folders and themes, as well as in the exploration of relationships and patterns using NVivo’s query and visualization tools. A coding tree will be developed through NVivo (40). A narrative summary will accompany any tabulated or charted results and will describe how the results relate to the review’s objective and questions.

Following data extraction, qualitative findings from various studies will be systematically organized, and recurrent themes will be identified. Themes will be analyzed to discern commonalities, variations, and nuanced perspectives across the qualitative studies. This process will facilitate a comprehensive understanding of the qualitative evidence regarding the research question. Knowledge gaps identified in the literature found through application of this protocol will also be presented. Quantitative data extracted from eligible studies will be synthesized, including frequencies, means, and measures of association. An integrated approach to data analysis will be used for mixed methods studies, in which qualitative and quantitative findings will be analyzed separately and then integrated to provide a holistic understanding.

#### Synthesis across methodologies

Following the individual analyses of qualitative, quantitative, and mixed methods studies, an overarching synthesis will be conducted to integrate findings across studies. Overarching themes, patterns and trend that emerge from the collective body of evidence will be identified. This will be accomplished through coding of results across methodologies into larger themes, when appropriate. Whenever possible, data will be presented in graphical or tabular from, to aid in understanding the research question, study participant and PRD specifics, facilitators of and barriers to DHL, and outcomes of participants with more facilitators to or barriers of DHL. Any statistical analyses will be limited to descriptive statistics and not result in a synthesis of the results or outcomes (30). The synthesis will provide a comprehensive and nuanced understanding of the barriers and facilitator to DHL in children and adolescents with PRD.

## Discussion

The objective of this ScR protocol is to provide a framework that will allow for the systematic identification of the facilitators of and barriers to DHL in children and adolescents with PRD and/or the primary caregivers of children or adolescents with PRD identified in the literature. This protocol will enable the examination of the intersection of technology, healthcare and the unique challenges endured by children with PRD. This ScR protocol will enable other researchers to plan and conduct reviews on the topics of DHL and digital health in the context of PRD and could be adapted for application to other chronic pediatric diseases.

### Potential limitations

Potential limitations of this ScR protocol may arise from the specified search terms, potentially resulting in incomplete literature coverage due to limitations in databases, language restrictions or evolving terminology may miss certain studies. The terminology related to DHL, facilitators to DHL, and barriers to DHL has not yet been standardized or universally accepted. This is evident in the array of keywords used in the pilot search of this protocol (Table 2) for concepts associated with DHL and digital health. The absence of consensus may pose challenges in locating pertinent articles.

Although we have attempted to be thorough and exhaustive, the search strategy outlined in this protocol may not capture all the published records on this subject. The search terms will be updated as new information emerges, if required. As this ScR protocol does not include searching the grey literature, it is possible that relevant information in non-journal format may be missed. Likewise, limiting the results of applying this protocol to articles in English only may also miss relevant information.

While multiple ScR frameworks are available (30,36), the JBI ScR approach (30) was selected because of its comprehensiveness, high levels of rigor, and widespread adoption. This framework, however, has been critiqued for lacking detailed guidance on certain aspects of the methodology, such as inclusion criteria and presentation of results (41).

The MMAT (38) is designed to appraise multiple types of studies (qualitative, quantitative and mixed methods studies), and thus lacks the depth of appraisal that may be available through tools such as the CASP (Critical Appraisal Skills Programme) suite of tools (42). However, the MMAT is a comprehensive instrument that uses a wide range of criteria and comes with clear and complete instructions.

## Data Availability

All data produced in the present work are contained in the manuscript and Appendix file

## Abbreviations

DHL: digital health literacy
ScR: scoping review
PRISMA-ScR: Preferred Reporting Items for Systematic Review and Meta-Analysis extension for scoping reviews
mHealth: mobile health
PRD: pediatric rheumatic diseases
JIA: juvenile idiopathic arthritis
MeSH: medical subject headings
PRESS: Peer Review of Electronic Search Strategies
MMAT: Mixed Methods Appraisal Tool

## Acknowledgements

The authors wish to acknowledge Caitlin Bakker from the Dr. John Archer Library and Archives, University of Regina, Saskatchewan, Canada for her PRESS review of the search terms.

## Ethics approval and consent to participate

Not applicable.

## Consent for publication

Not applicable.

## Availability of data and materials

All data generated or analyzed during this study are included in this published article.

## Authors’ contributions

CE drafted the initial protocol with input from DG, AR, JS, and MC. MC developed and refined the search strategies and the inclusion and exclusion criteria in collaboration with DG and CE. All authors were involved in reviewing and revising the article.

## Author information

## References

1. Competition Bureau Canada. Empowering health care providers in the digital era: digital health care market study, part 3 [Internet]. Gatineau QC: Competition Bureau Canada = Bureau de la concurrence Canada; 2022. (Empowering health care providers in the digital era). Available from: https://ised-isde.canada.ca/site/competition-bureau-canada/en/how-we-foster-competition/education-and-outreach/publications/Empowering-health-care-providers-in-the-digital-era

2. Norgaard O, Furstrand D, Klokker L, Karnoe A, Batterham R, Kayser L, et al. The e-health literacy framework: A conceptual framework for characterizing e-health users and their interaction with e-health systems. Knowl Manag E-Learn. 2015;7(4):522–40.

3. Laxer RM, Cellucci T, Rozenblyum E, editors. A Resident’s Guide to Pediatric Rhematology. 4th ed. The Hospital For Sick Children; 2019. 115 p.

4. Butler S, Sculley D, Santos D, Fellas A, Gironès X, Singh-Grewal D, et al. Effectiveness of eHealth and mHealth Interventions Supporting Children and Young People Living With Juvenile Idiopathic Arthritis: Systematic Review and Meta-analysis. J Med Internet Res. 2022 Feb 2;24(2):e30457.

5. Taylor ML, Thomas EE, Vitangcol K, Marx W, Campbell KL, Caffery LJ, et al. Digital health experiences reported in chronic disease management: An umbrella review of qualitative studies. J Telemed Telecare. 2022 Dec;28(10):705–17.

6. Faux-Nightingale A, Philp F, Chadwick D, Singh B, Pandyan A. Available tools to evaluate digital health literacy and engagement with eHealth resources: A scoping review. Heliyon. 2022 Aug;8(8):e10380.

7. Dadaczynski K, Okan O, Messer M, Leung AYM, Rosário R, Darlington E, et al. Digital Health Literacy and Web-Based Information-Seeking Behaviors of University Students in Germany During the COVID-19 Pandemic: Cross-sectional Survey Study. J Med Internet Res. 2021 Jan 15;23(1):e24097.

8. Norman CD, Skinner HA. eHEALS: The eHealth Literacy Scale. J Med Internet Res. 2006 Nov 14;8(4):e27.

9. van Kessel R, Wong BLH, Clemens T, Brand H. Digital health literacy as a super determinant of health: More than simply the sum of its parts. Internet Interv. 2022 Mar;27:100500.

10. Estrela M, Semedo G, Roque F, Ferreira PL, Herdeiro MT. Sociodemographic determinants of digital health literacy: A systematic review and meta-analysis. Int J Med Inf. 2023 Sep;177:105124.

11. Refahi H, Klein M, Feigerlova E. e-Health Literacy Skills in People with Chronic Diseases and What Do the Measurements Tell Us: A Scoping Review. Telemed E-Health. 2023 Feb 1;29(2):198–208.

12. Slater H, Stinson JN, Jordan JE, Chua J, Low B, Lalloo C, et al. Evaluation of Digital Technologies Tailored to Support Young People’s Self-Management of Musculoskeletal Pain: Mixed Methods Study. J Med Internet Res. 2020 Jun 5;22(6):e18315.

13. Choukou MA, Sanchez-Ramirez DC, Pol M, Uddin M, Monnin C, Syed-Abdul S. COVID-19 infodemic and digital health literacy in vulnerable populations: A scoping review. Digit Health. 2022 Jan;8:205520762210769.

14. World Health Organization. Global strategy on digital health 2020-2025 [Internet]. Geneva; 2021. Available from: https://www.who.int/docs/default-source/documents/gs4dhdaa2a9f352b0445bafbc79ca799dce4d.pdf

15. Buyting R, Melville S, Chatur H, White CW, Légaré JF, Lutchmedial S, et al. Virtual Care With Digital Technologies for Rural Canadians Living With Cardiovascular Disease. CJC Open. 2022 Feb;4(2):133–47.

16. Knapp C, Madden V, Wang H, Sloyer P, Shenkman E. Internet Use and eHealth Literacy of Low-Income Parents Whose Children Have Special Health Care Needs. J Med Internet Res. 2011 Sep 29;13(3):e75.

17. Wilson J, Heinsch M, Betts D, Booth D, Kay-Lambkin F. Barriers and facilitators to the use of e-health by older adults: a scoping review. BMC Public Health. 2021 Dec;21(1):1556.

18. Rosário R, Martins MRO, Augusto C, Silva MJ, Martins S, Duarte A, et al. Associations between COVID-19-Related Digital Health Literacy and Online Information-Seeking Behavior among Portuguese University Students. Int J Environ Res Public Health. 2020 Dec 2;17(23):8987.

19. Tschamper MK, Wahl AK, Hermansen Å, Jakobsen R, Larsen MH. Parents of children with epilepsy: Characteristics associated with high and low levels of health literacy. Epilepsy Behav. 2022 May;130:108658.

20. Knapp C, Madden V, Marcu M, Wang H, Curtis C, Sloyer P, et al. Information seeking behaviors of parents whose children have life-threatening illnesses: Information Seeking in Pediatric Palliative Care. Pediatr Blood Cancer. 2011 May;56(5):805–11.

21. Ditzler N, Greenhawt M. Influence of health literacy and trust in online information on food allergy quality of life and self-efficacy. Ann Allergy Asthma Immunol. 2016 Sep;117(3):258-263.e1.

22. Dağhan Ş, Kalkim A, Unlu E, Sahin HK, Yuksel M. Factors Affecting eHealth Literacy of Early Adolescents: School-based Research. Compr Child Adolesc Nurs. 2022 Oct 2;45(4):383–94.

23. Hider S, Muller S, Gray L, Manning F, Brooks M, Heining D, et al. Digital exclusion as a potential cause of inequalities in access to care: a survey in people with inflammatory rheumatic diseases. Rheumatol Adv Pract. 2022 Dec 30;7(1):rkac109.

24. Harris K, Jacobs G, Reeder J. Health Systems and Adult Basic Education: A Critical Partnership in Supporting Digital Health Literacy. HLRP Health Lit Res Pract [Internet]. 2019 Jul [cited 2021 Jul 6];3(3). Available from: http://journals.healio.com/doi/10.3928/24748307-20190325-02

25. Jafree SR, Bukhari N, Muzamill A, Tasneem F, Fischer F. Digital health literacy intervention to support maternal, child and family health in primary healthcare settings of Pakistan during the age of coronavirus: study protocol for a randomised controlled trial. BMJ Open. 2021 Mar;11(3):e045163.

26. Smith B, Magnani JW. New technologies, new disparities: The intersection of electronic health and digital health literacy. Int J Cardiol. 2019 Oct;292:280–2.

27. Juvalta S, Kerry MJ, Jaks R, Baumann I, Dratva J. Electronic Health Literacy in Swiss-German Parents: Cross-Sectional Study of eHealth Literacy Scale Unidimensionality. J Med Internet Res. 2020 Mar 13;22(3):e14492.

28. Manganello JA, Falisi AL, Roberts KJ, Smith KC, McKenzie LB. Pediatric injury information seeking for mothers with young children: The role of health literacy and ehealth literacy. J Commun Healthc. 2016 Jul 2;9(3):223–31.

29. Arksey H, O’Malley L. Scoping studies: towards a methodological framework. Int J Soc Res Methodol. 2005 Feb;8(1):19–32.

30. Peters M, Godfrey C, McInerney P, Munn Z, Trico A, Khalil H. Chapter 11: Scoping Reviews. In: Aromataris E, Munn Z, editors. JBI Manual for Evidence Synthesis [Internet]. JBI; 2020 [cited 2021 Jul 27]. Available from: https://wiki.jbi.global/display/MANUAL/Chapter+11%3A+Scoping+reviews

31. Tricco AC, Lillie E, Zarin W, O’Brien KK, Colquhoun H, Levac D, et al. PRISMA Extension for Scoping Reviews (PRISMA-ScR): Checklist and Explanation. Ann Intern Med. 2018 Oct 2;169(7):467–73.

32. Veritas Health Innovation, Ltd. Covidence [Internet]. Melbourne, Austrialia: Veritas Health Innovation, Ltd.; 2024. Available from: https://www.covidence.org

33. Lumivero, LLC. NVivo [Internet]. Lumivero, LLC; 2022. Available from: https://lumivero.com/products/nvivo/

34. McGowan J, Sampson M, Salzwedel DM, Cogo E, Foerster V, Lefebvre C. PRESS Peer Review of Electronic Search Strategies: 2015 Guideline Statement. J Clin Epidemiol. 2016 Jul;75:40–6.

35. Parnell TA. Health Literacy: History, Definitions, and Models. In: Health Literacy in Nursing [Internet]. New York: Springer Publishing Company; 2014. p. 3–32. Available from: https://connect.springerpub.com/content/book/978-0-8261-6173-4/part/part01/chapter/ch01

36. Peters MDJ, Marnie C, Colquhoun H, Garritty CM, Hempel S, Horsley T, et al. Scoping reviews: reinforcing and advancing the methodology and application. Syst Rev. 2021 Dec;10(1):263.

37. Page MJ, McKenzie JE, Bossuyt PM, Boutron I, Hoffmann TC, Mulrow CD, et al. The PRISMA 2020 statement: an updated guideline for reporting systematic reviews. BMJ. 2021 Mar 29;372(n160):1–36.

38. Hong QN, Fàbregues S, Bartlett G, Boardman F, Cargo M, Dagenais P, et al. The Mixed Methods Appraisal Tool (MMAT) version 2018 for information professionals and researchers. Educ Inf. 2018 Dec 18;34(4):285–91.

39. Microsoft Corporation. Microsoft Excel. Redmond, WA: Microsoft Corporation; 2024.

40. Thompson Burdine J, Thorne S, Sandhu G. Interpretive description: A flexible qualitative methodology for medical education research. Med Educ. 2021 Mar;55(3):336–43.

41. Khalil H, Bennett M, Godfrey C, McInerney P, Munn Z, Peters M. Evaluation of the JBI scoping reviews methodology by current users. Int J Evid Based Healthc. 2020 Mar;18(1):95–100.

42. Critical Appraisal Skills Programme. CASP Checklists [Internet]. 2023. Available from: https://casp-uk.net/casp-tools-checklists/

