## Supplemental table for "Identifying Facilitators of and Barriers to Digital Health Literacy in Pediatric Rheumatic Diseases: A Scoping Review Protocol"

**Identifying Facilitators of and Barriers to Digital Health Literacy in Chronic Childhood Rheumatic Disease: A Scoping Review Protocol**

Craig Eling; Alan Rosenberg; Jennifer Stinson; Maryam Mehtar; Jasmin Bhawra; Mary Chipanshi; Roona Sinha; Donna Goodridge

**Appendix 1**

Table 1 presents facilitators of and barriers to DHL in children with or without a chronic disease, parents of children with or without a chronic disease, and adults with rheumatic disease.

| **Table 1.** Facilitators of and barriers to DHL in children with or without a chronic disease, parents of children with or without a chronic disease, and adults with rheumatic disease identified from the literature search. | |
| --- | --- |
| Facilitators of DHL | Desire and motivation to learn and use digital health services (16)  Positive past experiences with digital health and reliable sources of health information (17,18)  Belief that digital technologies and digital health is beneficial to personal health (13,17)  Practitioner recommendation and support for digital health (17)  Training/education (17)  Internet and/or devices accessible at public locations or through subsidization (17)  Parental education greater than high school (19–21) |
| Barriers to DHL | Lower socioeconomic status (4,16,19–23)  Limited English language proficiency (13,16,24)  Increasing rurality or geographic isolation (13,24,25)  Indigenous, immigrant, and marginalized populations (16,24,26)  Lack of access to broadband Internet, high-speed cellular networks, or lack of access to internet-capable and usable devices (6,17,23,24)  Aged 65-years or older (16,23,24) or too young to have developed digital skills (4) |
